# Patients’ Experiences of Using Smartphone Apps to Manage their Gestational Diabetes Diagnosis: A Systematic Review - protocol (version 1)

**DOI:** 10.1101/2022.03.22.22272786

**Authors:** Ali Anis, Annette Plüddemann, Anurag Choksey, Clara Portwood, Vikram Mitra, Carl Heneghan

## Abstract

**Background:** Gestational diabetes is a strong predictor of type 2 diabetes onset. However, women who make healthy dietary and lifestyle choices can significantly reduce their risk. There is a need to understand the factors facilitating or preventing women from using smartphone apps that may encourage these behaviours.

**Methods:** We plan to conduct a systematic review of patient experiences when using mobile health applications to manage gestational diabetes. We will include primary studies of qualitative data around patient experiences of using these applications, as well as the barriers and facilitators to using technologies. We will search the following electronic databases: Medline, Embase, PsycINFO, Global Health, Web of Science, Cochrane Central Register of Controlled Trials (CENTRAL), AMED and CINAHL and manually search the reference lists of included studies. We will include primary studies involving direct user interviews and where the results have been analysed using qualitative methods. To assess the quality of included studies, we will use the CASP qualitative research checklist.

**Expected results:** We intend to use summary tables to report the characteristics of the study population, the overarching themes that emerged, and recommendations for research and practice.

## Background

Gestational diabetes mellitus (GDM), which describes glucose intolerance first recognized during pregnancy [1], is the most common medical complication of pregnancy [2]. As the single strongest population predictor of type 2 diabetes onset [3, 4]; it is estimated that up to 50% of mothers develop overt diabetes within 10 years of their initial GDM diagnosis [5]. In high-risk populations, however, it has been demonstrated that lifestyle interventions can reduce the incidence of type 2 diabetes by 58% [6].

Increasingly, smartphone-based health applications are used by patients to manage all aspects of their health [7]. A recent cross-sectional study estimated that in 2017 more than half of all global smartphone users had health applications installed on their device [7]. The number of applications dedicated to diabetes surpasses those available for any other prevalent health condition [8], and such apps have been shown to positively affect health behaviours including physical activity and diet [9]. Furthermore, it has also been reported that mobile apps may improve glycemic control markers specifically in the context of GDM patients [10].

Therefore, we plan to perform a systematic review of qualitative data on patient experiences of using mobile apps to manage their GDM diagnosis. We will produce a meta-synthesis of themes for the factors stimulating and preventing patients from using these technologies. The results of our review could be used to guide practice and inform research on the development of more effective applications.

## Methods

### Objectives

#### The specific focus of the review

- Population: women with an existing or recent diagnosis of gestational diabetes
- Intervention: any mobile health application to manage diabetes
- Outcomes: patient experiences, barriers and facilitators to app usage, improvement suggestions

#### Type of studies

We will include primary studies that feature qualitative data on GDM patients’ experiences with using mobile phone applications.

#### Search strategy

We will search the following electronic databases: Medline, Embase, PsycINFO, Global Health, Web of Science, Cochrane Central Register of Controlled Trials (CENTRAL), AMED and CINAHL from Jan 2010 onwards and manually search the reference lists of included studies. The searches will include term variations of mobile applications (mobile app* OR smartphone app* OR portable electronic app*) and gestational diabetes mellitus and related terms. Two authors will independently screen all titles and abstracts for inclusion. The need for additional search terms will also be reviewed at this stage. The final search strategy will be developed with advice from information specialists and will be included in the appendix of our manuscript for publication. The full texts of potentially eligible studies will be assessed independently in duplicate by two study authors. Disagreements will be resolved by discussion or by consulting a third review author.

#### Eligibility criteria

We will include primary studies of qualitative data on GDM patient experiences with using mobile apps to manage GDM. Additionally, the results must be derived from direct user interviews held either in person, by telephone or via video conference, or they may be taken from focus groups. We will exclude studies that use only surveys to assess patient experiences. Reviews, commentaries and editorials will be excluded, but reference lists will be searched for any additional studies that meet the inclusion criteria.

#### Types of intervention

Any mobile health application to manage diabetes

#### Quality assessment

Quality will be assessed and recorded by one study author using the CASP qualitative research checklist [11]. A second reviewer will then independently check the scoring. Disagreements will be resolved by discussion or by consulting a third review author. We will also assign a judgement about the overall quality of each study: ‘low’, ‘high’ or ‘unclear’ risk of bias, based on how the study fared against each of the ten items in the CASP checklist.

#### Data extraction

We will adhere to the PRISMA reporting guidelines [12] and make use of a flow diagram to report on the number of studies included and excluded at each stage. Studies which are excluded at the full text screening stage will be tabulated alongside reasons for exclusion.

One study author will perform data extraction and quality assessment of the included studies, and this will be independently checked by a second author. We will extract data on the population, study characteristics (e.g. how many interviews were conducted, the method of data collection, etc.), the intervention (i.e. the mobile app used) and the outcomes of interest.

#### Outcomes of interest

We will prioritise outcomes that can guide practice or can inform research on the development of new mobile apps. User experiences, patient-reported barriers and facilitators of app usage, and improvements suggested by direct users will be considered.

We will perform a meta-synthesis of the major themes that emerge and will use summary tables to present the evidence. We will report the frequency with which the individual themes arose and offer examples of quotations from patients that strongly reflect the themes that are identified. Alongside any improvements suggested by patients, these themes will be used to outline clear recommendations for research and practice.

## Data Availability

All data produced in the present study will be available upon reasonable request to the authors

## Authors’ contributions

AA devised the topic for research, wrote the first draft and edited this version. AP and CH helped to refine the study question and eligibility criteria. All authors contributed to subsequent drafts and approved the final version.

## Ethics committee approval

No ethics approval was necessary.

## Data Availability

All data included in the review will be provided in the tables and text.

